# Mapping the common and rare variant genetic risk landscape for pulmonary fibrosis

**DOI:** 10.64898/2026.05.15.26351995

**Authors:** SEM Lucas, K Raspin, NE Nelson, PS Graham, I Cox, S Chear, C Zappala, G Keir, N Goh, P Hopkins, SJ Ellis, V Navaratnam, W Cooper, I Glaspole, PN Reynolds, C Chia, C Grainge, P Kendall, L Troy, N Nunez Martinez, AL Peljto, TA Fingerlin, DA Schwartz, SLF Walsh, Y Moodley, EH Walters, J Robertson, MR Emerson, M Joy, SB Cohen, TM Bryan, D Chambers, JA Mackintosh, T Corte, JL Dickinson

**Affiliations:** Menzies Institute for Medical Research, University of Tasmania, Australia; Royal Brisbane and Women’s Hospital, Queensland, Australia; Princess Alexander Hopsital, Brisbane, Queensland, Australia; The Alfred Hospital, Melbourne, Victoria, Australia; Institute for Breathing and Sleep, Melbourne, Australia; University of Melbourne, Melbourne, Australia; University of Queensland, Queensland, Australia; Department of Thoracic Medicine, The Prince Charles Hospital, Brisbane Queensland, Australia; Royal Prince Alfred Hospital, Sydney, New South Wales, Australia; Royal Adelaide Hospital, Adelaide, South Australia, Australia; Launceston General Hospital, Tasmania Australia; John Hunter Hospital, Newcastle, New South Wales, Australia; Respiratory Medicine Service, Albany, WA, Australia; School of Medicine and Pharmacology, University of Western Australia, Perth, WA, Australia; University of Sydney, New South Wales, Australia; Department of Medicine, Division of Pulmonary Medicine, University of Colorado, Colorado; The National Jewish Health Cohen Family Asthma Institute, Division of Allergy and Immunology, National Jewish Health, Denver, Colorado; Qureight Ltd, Cambridge, UK; National Heart and Lung Institute, Imperial College London, UK; Royal Perth Hospital, Western Australia, Australia; Tasmanian School of Medicine, University of Tasmania; Border Physicians Group, West Albury, NSW, Australia; Children’s Medical Research Institute, Faculty of Medicine and Health, University of Sydney, Westmead, New South Wales, Australia; Queensland Lung Transplant Service, The Prince Charles Hospital, Chermside, Australia; Faculty of Health, Medicine and Behavioural Sciences, The University of Queensland, Australia; Australian Centre for Cellular Aging (ACCA), The Prince Charles Hospital, Chermside, Australia; Faculty of Medicine, University of Queensland, Brisbane, Australia; Monash University, Melbourne, Australia

## Abstract

**Background:** Genetic studies to date are yet to define the major portion of the genetic risk for adult-onset pulmonary fibrosis (PF). Further, the dearth of knowledge of clinically actionable variants for PF is hampering efforts to implement genetic testing to aid early diagnosis and improve disease management. Here we evaluated the contribution of rare and common variants to PF in cohorts with and without a family history of PF.

**Method:** Whole genome sequencing (WGS) was performed in a familial cohort comprising PF cases and their family members (85 individuals representing 55 families); and 122 cases from the Australian IPF Registry (AIPFR) with and without a self-reported family history of PF. WGS data were interrogated for rare potentially PF-causing variants in 33 genes previously associated with PF. Variants that were rare and predicted to be likely causative were formally curated using the American College of Medical Genetics and Association for Molecular Pathology (ACMG-AMP) guidelines. Additionally, to examine the common genetic risk variant contribution, a weighted polygenic risk score (PRS) was generated using 16 confirmed IPF-associated common variants. PRS were generated from WGS for the 85 clinically confirmed familial cases and 122 AIPFR cases. In the remaining 202 AIPFR cases, PRS were generated from TaqMan genotyping data.

**Results:** Interrogation of WGS generated from 207 individuals with PF revealed multiple rare putative pathogenic variants in the familial and non-familial cohorts. Formal curation of the familial cohort revealed segregating pathogenic (P) or likely pathogenic (LP) variants in *TERT* or *RTEL1* in four families (7.3%) with the majority of remaining variants classified as variants of uncertain significance (VUS; 12.7%) in seven additional families. A further analysis of non-segregating *TERT* variants in families with at least two recruited relatives with PF revealed two additional LP variants (8% of multi-case families) and a VUS. Amongst non-familial participants, four variants met the threshold for classification as P/LP variants (3.3%), with a further six individuals found to harbour VUS following curation (4.9%). Overall weighted PRS did not differ significantly between individuals with familial PF or with no reported family history. However, PRS in all patient groups were significantly elevated compared with population controls.

**Conclusion:** A rare LP/P variant was identified 9.1% of families and individuals with clinically confirmed familial PF and 3.3% of apparently non-familial PF. This included a non-segregating LP variant in *TERT* that highlights the complexity of PF inheritance. VUS remain the most frequent category of rare variants identified in known PF-related genes. For ∼75% of individuals with a confirmed family history no potentially causative variants were identified in known PF-related genes, nor was there evidence that a high burden of common variants contributed to risk in these families. Similarly, we found no evidence that a high burden of common variants contributes to a significant proportion of risk PF in individuals without a reported family history.

## INTRODUCTION

Pulmonary fibrosis (PF) is a devastating lung disease leading to impaired gas exchange, declining lung function and ultimately respiratory failure. Idiopathic Pulmonary Fibrosis (IPF) is the most common and severe form, representing >30% of cases^1^, and consequently the most well studied. Originally thought of as a condition caused by smoking and other environmental risks, we now understand that both genetic and environmental factors underpin the aetiology of this disease. Genetic studies of familial PF have identified key biological pathways and genes that predispose individuals and families to PF. Indeed, family history is a one of the strongest risk factors for IPF^2^, as well as other forms of PF. This is further highlighted by the observation that different PF subtypes occur within families^3^, with different PF presentations occurring within members of the same family who carry the same rare disease-causing variant^4^. Recent studies have highlighted that familial PF is self-reported in 20-25% of individuals with IPF and 12.5-15% of other subtypes of PF^5, 6^. A prospective study of individuals with apparently sporadic IPF reported 33% were reclassified as familial PF when first degree relatives over the age of 40 were screened with high resolution CT scans, with over 50% of siblings found to have PF or preclinical disease (interstitial lung abnormalities)^7^. Subsequently these findings were supported in a second study where preclinical or established PF was identified in 36% of relatives of individuals with apparently sporadic IPF^8^. In familial PF, 16%^9^ to 46%^10^ of relatives were found to have radiological evidence of preclinical or established PF. Even based on the lowest frequency of undiagnosed disease in relatives, the incidence of preclinical disease is 100 times the incidence of IPF in the general population^9^.

The first rare PF-causing variant was discovered in the surfactant gene, *SFTPC*, in 2001^11^ and since this discovery there has been a rapid expansion in our knowledge of high risk genes causative of PF. This includes the identification of causative variants in other surfactant genes, *SFTPA1* and *SFTPA2*^12^, and genes involved in essential processes for type II alveolar epithelial cells such as vesicular trafficking. However, the majority of rare PF-causing variants are in telomere biology genes. In fact, over 90% of individuals with PF with a known rare disease-causing variant are located in just five genes, all telomere biology genes: *TERT*, *TERC*, *RTEL1*, *PARN* and *DKC1*^13^. Individuals with a telomere biology disorder (TBD) frequently present with PF but may also develop bone marrow failure, haematological malignancy and liver disease, and often multiple presentations may be observed within the same family^14, 15^. In addition, to telomere biology associated genes,

Multiple genome wide association studies (GWAS) have been published for IPF since the first in 2013^16^, with the largest meta-analysis recently identifying 34 independent associations with evidence of replicability at 23 loci associations^17^. Notably, many of the associated variants are located in, or close to, genes residing in the same biological pathways as the rare PF-causing genes, most particularly the telomere biology genes *TERT*, *TERC* and *RTEL1*. Interestingly, the strongest association and the largest effect size is imparted by a single gain of function variant in the promoter of *MUC5B* (rs35705950) which is associated with an approximate 5-fold increase in risk of developing PF for heterozygous individuals^18^. Other common variants have also been identified, with a smaller impact on risk, typically with odds ratios (OR) of approximately 1.5. Several of these variants highlight inflammatory processes due to their proximity to genes involved in these biological pathways (*TGFB1, IL1RA, IL8, TLR3, FAM13A, TOLLIP)*. While these findings have not yet been translated into the clinic, there is increasing interest in utilising these data to develop polygenic risk scores (PRSs) that may predict individuals at risk of PF development^19^.

Traditionally, rare variants were thought to only contribute to familial PF, while common variants have been considered key genetic drivers in apparently sporadic disease. However, recent studies have highlighted that rare PF-causing variants can be identified in approximately 10% of individuals without a family history of disease^20–22^. Therefore, this study sought to explore the contribution of rare variants in an Australian cohort in individuals with familial and non-familial PF. Additionally, this study aimed to explore the common variant contribution to PF in both groups to gain a better understanding of the genetic underpinnings of PF.

## METHODS

### Participants

#### Individuals with PF (Australian IPF Registry cases)

The Australian IPF Registry (AIPFR) recruited individuals with IPF from across Australia, as outlined previously^23^. Briefly, participants completed questionnaires every six months gathering demographic information, smoking history and medical history. A subset of participants provided blood samples with DNA available for 324 cases. Of the 324 cases included in this study, 41 self-reported a family history of PF with the remainder of AIPFR termed “non-familial IPF” (n = 284). This study received ethical approval from the Sydney Local Health District approved protocols X11-0287 and X14-0264.

#### Individuals with PF (Genetic Research in Pulmonary Fibrosis; GRIPF cases)

The GRIPF cohort comprises individuals diagnosed with PF with a positive family history and their family members where available, and recruitment has been previously described^10^. Informed consent and a physical examination were conducted by a respiratory physician and participants completed questionnaires, a chest high resolution computed tomography (HRCT) scan, full pulmonary function testing (PFT), and saliva or blood collection for genetic analysis. As part of the GRIPF study, any additional affected relatives and at least one self-reported unaffected relative were also invited to participate, where available. This cohort comprised clinically confirmed familial PF (n = 85 from 55 families), including five families with three relatives with PF, 20 families with two relatives with PF cases and 30 probands without additional recruited affected relatives. The study received ethical approval from the Sydney Local Health District (X18-0193 & 2019/ETH06444) and Project ID: 53369; HREC/2019/QPCH/53369.

A summary of the number of participants included in each analysis for each PF group is provided in Figure 1.

**Figure 1.**
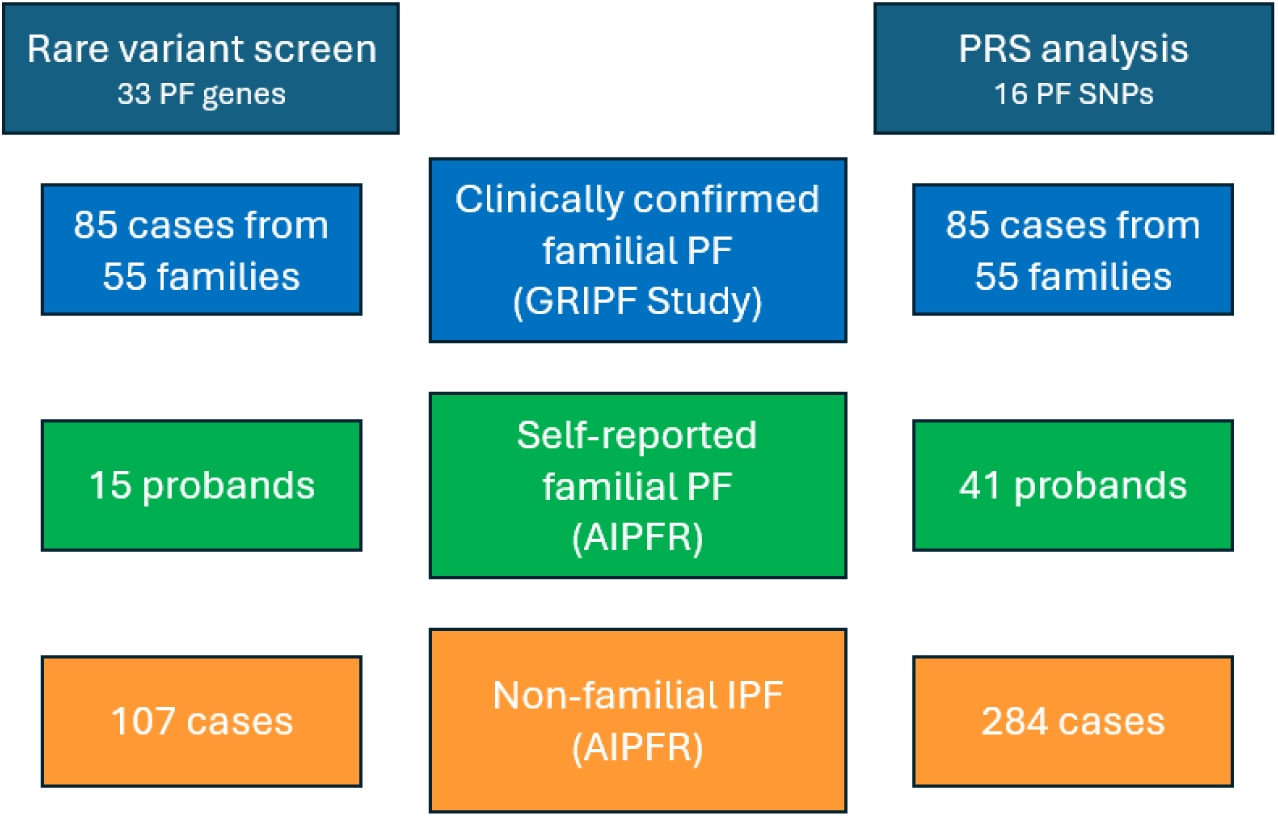
PF groups included in analysis

#### Population Controls (1000 Genomes Project)

Population control data from 404 unrelated non-Finnish Europeans available as part of the 1000 Genomes Project^24^ were utilised for the common variant analysis. This included whole genome sequencing (WGS) data for the following populations: British in England and Scotland, Iberian populations in Spain, Toscani in Italy, and Utah residents (CEPH) with Northern and Western European ancestry.

### DNA extraction for individuals with PF

For participants participating in the GRIPF Study, genomic DNA was extracted from peripheral blood samples using the Nucleon BACC Genomic DNA Extraction Kit (Cytiva Life Sciences). Blood samples for the AIPFR Participants were processed and stored as buffy coats in liquid nitrogen until extraction with the QiaAmp DNA Mini Kit (Qiagen).

### Genetic Resources

#### Whole genome sequencing

Whole-genome sequencing (WGS) data was available for 85 individuals with familial PF from 55 families recruited to the GRIPF Study. These data were generated using PCR-free library construction with 150bp paired-end sequencing on an Illumina NovaSeq6000 (Australian Genome Research Facility) or TruSeq PCR-free libraries with 150bp paired-end sequencing on the NovaSeq6000 (Macrogen Oceania). FASTQ files were provided and processed in-house using our high-performance computing genomics cluster, Rosalind. Using the Sarek^25^ v3.0.1, reads were aligned to the GRCh38 reference genome with Burrows-Wheeler Aligner^26^ v0.7.17-r1188 and single nucleotide variants (SNVs) and small insertions and deletions (indels) were joint-called using the Genome Analysis Toolkit Best Practices pipeline^27^.

WGS data was available for 122 individuals from the AIPFR registry, including 15 individuals that reported a family history of PF. These data were generated using PCR-free library construction sequenced with 30x coverage with Illumina HiSeq X technology performed by the NIH Trans-Omics for Precision Medicine (TOPMed) Program^28^, as described previously^29^.

#### Genotyping for common IPF variants

For the additional AIPFR participants, for which WGS was not available, 16 single nucleotide polymorphisms (SNPs) were selected for genotyping based on a significant association with IPF in the genome-wide association study (GWAS) by Allen *et al*.^30^. SNPs were genotyped either by direct sequencing using primers obtained from Integrated DNA Technologies and sequenced on the ABI3000 or end-point genotyping using TaqMan probes obtained from Thermo Fisher Scientific and performed on the Roche LightCycler 480.

### Genetic analysis

#### Relationship testing

For the individuals with PF with WGS data available, relationship testing was conducted to confirm that families were unrelated to each other and validate the reported familial relationships. Identify-by-decent estimates were calculated in Plink v1.9^31^ using 263,547 linkage disequilibrium pruned autosomal SNPs based on the Illumina Infinium OmniExpress array. These data were visualised in R using Plotly^32^. A single AIPFR participant who reported a family history was determined to be an affected brother of the proband from the GRIPF Study and thus was moved to the clinically confirmed familial PF group for analysis.

#### Polygenic risk score generation and analysis

To investigate the contribution of common genetic variation to IPF, a polygenic risk score (PRS) was calculated using the 16 SNPs that reached significance in Allen *et al*.^30^, weighting the sum of the risk alleles by their respective effect sizes. The included SNPs and the reported odds ratios used for weighting are outlined in **Table S1**.

For the clinically confirmed familial cohort (GRIPF) and population controls, the SNPs of interest were extracted from the WGS data using BCFtools^33^, whereas the genotyping data generated using direct sequencing or end-point genotyping data was utilised for the AIPFR participants (both the self-reported familial and non-familial cases). The distribution of PRS scores for each group were plotted in R using ggplot2. Statistical analysis was conducted in R^34^ version 4.1.0 using a one-way ANOVA, and post hoc testing used a Tukey HSD test.

### Rare variant gene screen and variant curation

A total of 33 candidate genes were selected for inclusion, comprising genes with published evidence for rare variant causation in familial PF cases, or syndromes of which PF is a feature, for example TBD (**Table S2**). Variants within these genes were annotated with refGene, avsnp150 and minor allele frequencies from gnomAD v2.1.1 using ANNOVAR^35^ (v20191024). CADD v1.6^36^ scores were obtained via an online batch submission (https://cadd.gs.washington.edu/score).

Qualifying variants identified in individuals with PF were exonic variants (excluding UTRs) and splice-site variants that were rare (MAF < 0.01) in all populations of gnomAD and had a CADD score of at least 15. For all PF groups, heterozygous variants located in genes known to cause disease in an autosomal recessive manner, without a second qualifying variant located within the same gene or related gene (e.g. *ABCA3* and a surfactant gene), were excluded. In the familial cohort, segregation of qualifying variants was additionally considered on a per family basis and non-segregating variants were excluded, noting that this was not the case for *TERT* variants. Due to previous published findings^37^, the potential role of non-segregating variants in *TERT* in disease causation was further explored and these variants were additionally curated. Included variants were curated using the ACMG-AMP guidelines^38^. Specific rules are outlined in **Table S3**. Rules for *TERT* variants were based on those previously published^39^, with some minor adjustments largely to account for the release of gnomAD 4.1.1.

### Functional assays for *TERT* variants

Telomerase activity and processivity was determined using a direct assay for *TERT* variants where functional evidence would aid variant curation and interpretation. Each selected *TERT* variant was introduced by site-directed mutagenesis into a plasmid encoding FLAG-tagged hTERT under a CMV promoter. This plasmid was transfected into HEK293T cells along with a plasmid encoding *TERC* (telomerase RNA, component hTR) and *DKC1* (dyskerin), and hTERT protein levels were quantified by Western blot using an anti-FLAG antibody, normalised to tubulin as a loading control^40^. Wild-type (WT) *TERT* was transfected as a control. Telomerase was immunopurified using anti-FLAG M2 antibody affinity gel (Sigma), and the amount of hTR recovered after immunopurification of hTERT was quantified by northern dot-blot as described^40^. Equal volumes of immunopurified telomerase were used in direct telomerase activity assays, involving extension of a telomeric primer with radiolabeled nucleotides, followed by electrophoresis on a high-resolution acrylamide gel. The reactions were carried out as described^40^, except lower concentrations of DNA and nucleotide substrates were used to allow for greater sensitivity in detecting DNA and nucleotide binding defects (100 nM DNA oligonucleotide [TTAGGG]3, 15 μM dTTP, 10 μM dGTP, 10 μM dATP).

Telomerase activity (the enzyme’s ability to add nucleotides to a DNA molecule) was quantified by analysing the sample lanes and normalising the results to the amount of hTR recovered to give specific activity, expressed as a percentage of WT telomerase activity on the same gel. Processivity (the ability of telomerase to continuously extend a single DNA molecule) was quantified as previously described^41^.

## Results

The clinical characteristics for the PF groups are presented in **Table 1**. Overall, the clinically confirmed familial PF cases were younger, more likely to be female, less likely to smoke and have fewer pack years, if they did have a smoking history. For the familial cohort, all reported relationships were confirmed empirically, and no cryptic relatedness was identified between the reported families (**Figure S1**). Additionally, the clinically confirmed familial PF group had a significantly lower prevalence of pulmonary hypertension, heart disease, and diabetes, compared to the sporadic cohort. Allele counts for the *MUC5B* risk variant, rs35705950, are provided in **Table 1**. A total of 73.1% sporadic IPF Registry cohort and 78.8% of the familial PF cases harboured at least one risk allele, in keeping with previously published European population frequencies^42^.

**Table 1.**
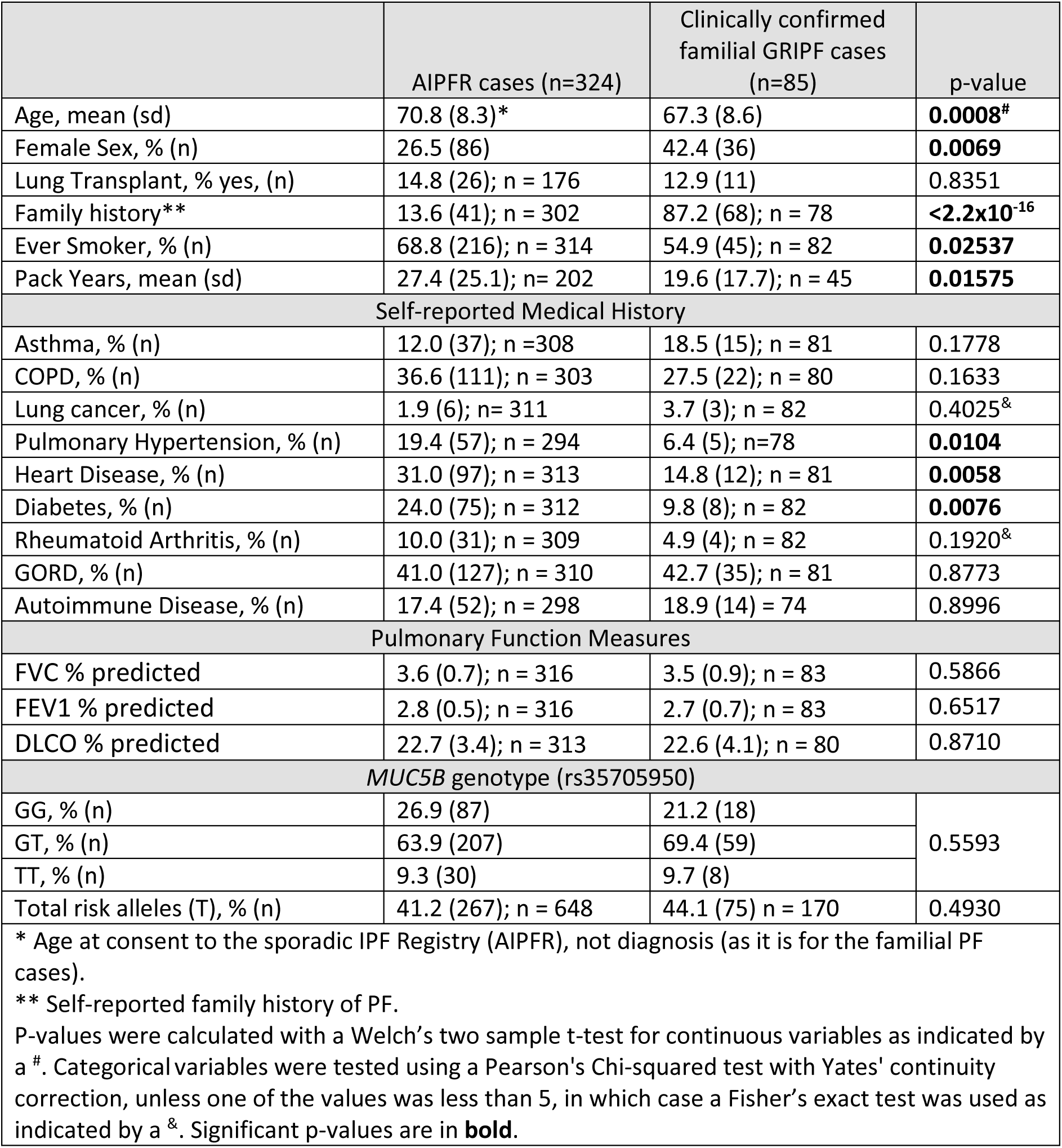
Clinical characteristics of the non-familial cases and cases from the familial cohort included in this study.

### Rare variants in known PF genes

#### Clinically confirmed familial PF cohort (GRIPF)

Utilising the rare variant prioritisation strategy outlined, 47 qualifying variants were identified in the clinically confirmed familial PF group. Utilising our candidate gene approach, 14 variants segregated with disease in family members for whom samples were available. Formal curation of these 14 variants is presented in **Table 2** and revealed three variants met the threshold for classification as pathogenic (P) and one variant was likely pathogenic (LP), with seven classified as variants of uncertain significance (VUS) and one variant was classified as likely benign (LB) and two as benign (B). This analysis revealed a genetic basis of disease in 7.3% of the families in the clinically confirmed familial cohort (4/55), while variants with some evidence for a role in disease causation were identified in a further 12.7% (7/55).

Of the remaining qualifying variants, 18 were located in genes associated with autosomal recessive disease, and as no individual harboured the required two variants in the same gene consistent with recessive disease, these variants were excluded from variant curation. This included the known likely pathogenic variant in *ABCA3* (p.R208W), which was observed in a single proband^43^, although it is acknowledged that a “second hit” in an alternate gene may be present in the affected pathway that has yet to be identified.

The remaining 15 qualifying variants were heterozygous and located in genes associated with autosomal dominant disease. Three of these were located in telomerase and were further investigated to explore the potential role of non-segregating variants in disease. Non-segregating variants in other genes were excluded for the purpose of this report. Direct sequencing of the three non-segregating variants in TERT revealed one as a sequencing artifact, however p.(R756H), p.(S602L) validated and were further investigated (**Table 3**). For both variants, two siblings with IPF were recruited, but only one of the siblings was found to carry the variant. The p.(R756H) variant was additionally observed in an unaffected niece of the recruited individuals who self-referred after the genomic sequencing was conducted for this study (via direct sequencing). The functional impacts of both p.(R756H) and p.(S602L) were assessed using our direct telomerase assay as described below. Variant curation classified the p.(R756H) variant as a VUS, however, the p.(S602L) variant was classified as LP and therefore was considered the genetic basis of disease in the family. This increased the identification rate in this cohort to 9.1% (5/55) of families. For the remaining families the genetic aetiology of disease remains unresolved.

#### AIPFR Cohort with no reported family history of PF

A total of 41 qualifying variants were identified in the non-familial cohort for whom whole genome sequencing was available (n=107). Eighteen variants across nine genes were curated as the remaining were excluded as only a single heterozygous variant was observed in a gene with an autosomal recessive inheritance pattern of disease. Five variants were heterozygous variants were in genes associated with autosomal recessive disease where the carriers were compound heterozygotes, however variant curation classified these variants as LB or VUS and therefore were not considered likely to contribute to disease in these individuals. Further discussion of this is presented in **Supplementary Section 4** and **Table S4**. A summary of the 13 remaining curated variants is presented in **Table 3**. One *TERT* variant met the threshold for classification as P and three *RTEL1* variants were LP; however, six variants remained classified as VUS. A further two met the criteria for a LB classification and one as B. The LP variant in *RTEL1,* p.(R1264H), that was also observed in an unrelated individual in the clinically confirmed familial PF cohort. Overall, a disease-causing variant was identified in 3.7% of the individuals in the non-familial cohort (n = 4/107) while an additional 7.5% carried a VUS (n = 8/107).

#### AIPFR Cohort with a reported family history

Eight qualifying variants were identified in the 15 individuals that self-reported familial PF where WGS data was available. Two variants were both present in *AP3B1* (NM_003664) and observed in the same individual. Following curation, the p.(Q1039K) variant was classified as a VUS and the p.(V999M) variant was LB, and therefore they were not considered causative for the disease in this individual (**Table S4**). A further six variants were subsequently excluded as they were heterozygous variants identified in a gene associated with autosomal recessive disease.

**Table 2.** Segregating variants identified in the clinically confirmed familial PF cohort.

| Gene | Position | Coding Change | Protein Change | gnomAD | CADD | REVEL | SpliceAI | ACMG-AMP | Ref |
| --- | --- | --- | --- | --- | --- | --- | --- | --- | --- |
| *TERT* | chr5:1253795 | c.3332 C>T | p.(T1111M) | 1.54 x 10<sup>-4</sup><br>(AAM) | 23.6 | 0.560 | 0.01 (AL/AG) | **VUS (4):** PS3_Mod, PP2, PP3 |  |
| *TERT* | chr5:1255413 | c.3033-2A>C | p.F1012Dfs*12 | abs | 33.0 | n/a | 0.99 (AL) | **P (10):** PVS1, PP4, PM2_Sup |  |
| *TERT* | chr5:1260529 | c.2915 G>A | p.R972H | 2.47x10<sup>-6</sup><br>(NFE) | 15.8 | 0.092 | 0.02 (DL) | **VUS (4):** PP2, PP4, PS3_Mod, PM2_Sup, BP4_Sup | 44-47 |
| *TERT* | chr5:1260593 | c.2851 C>T | p.R951W | 2.47x10<sup>-6</sup><br>(NFE) | 19.7 | 0.390 | 0.12 (AG) | **P (12):** PS4, PP1_Str, PS3_Mod, PP2, PP4, PM2_Sup, BP4_Sup | 4, 13, 48-50 |
| *TERT* | chr5:1266524 | c.2594 G>A | p.R865H | 1.24x10<sup>-6</sup><br>(NFE) | 26.3 | 0.939 | 0.01 (DL) | **P (12/14):** PS4, PS3_Mod, PP1_Mod, PP2, PP3, PP4, PM2_Sup | 4, 13, 20, 48, 49, 51 |
| *SFTPA2* | chr10:79559324 | c.160 C>T | p.P54S | 0 | 23.9 | 0.142 | 0.01 (AL) | **VUS (-1):** PM2_Sup, BP4_Mod |  |
| *NF1* | chr17:31159036 | c.231 A>T | p.K77N | 7.05x10<sup>-5</sup><br>(NFE) | 23.4 | 0.108 | 0.04 (AL) | **LB (-2):** BP4_Mod |  |
| *NF1* | chr17:31169939 | c.528 T>A | p.D176E | 4.63x10<sup>-3</sup><br>(NFE) | 21.8 | 0.641 | 0.01 (AG) | **B (SA):** BA1 |  |
| *RTEL1* | chr20:63662602 | c.452 A>G | p.K151R | abs | 23.5 | 0.079 | 0.63 (DG) | **VUS (2):** PP3, PM2_Sup |  |
| *RTEL1* | chr20:63685568-63685645 | c.1237_1266del | p.(A413_K422del) | abs | 27.0 | n/a | 0.89 (DL) | **VUS (4):** PVS1_Mod, PP4, PM2_Sup | 52* |
| *RTEL1* | chr20:63687670 | c.1381 G>A | p.(G461S) | 3.60 x10<sup>-5</sup> (SAS) | 23.6 | 0.567 | 0.01<br>(AL/DL/AG) | **VUS (1):** PP4 |  |
| *RTEL1* | chr20:63689838 | c.2114 A>G | p.(Y705C) | 7.90 x10<sup>-7</sup> (NFE) | 23.6 | 0.765 | 0.10 (AL) | **VUS (5):** PS4_Mod, PP3, PP4, PM2_Sup | 13 |
| *RTEL1* | chr20:63690835 | c.2444 G>T | p.(S815I) | 1.71x10<sup>-3</sup> (FIN) | 17.9 | 0.047 | 0.02 (AG) | **B (SA):** BA1, BP4_Mod | 52 |
| *RTEL1* | chr20:63695619 | c.3791 G>A | p.(R1264H) | 1.03x10<sup>-5</sup> (NFE) | 23.5 | 0.641 | 0.03 (DL) | **LP (7):** PS4, PP4, PM2_Sup PM3_Sup | 53-55 |

Positions are in hg38; coding and protein changes are based on the following transcripts: *TERT* (NM\_198253); *SFTPA2* (NM\_001320813); *NF1* (NM\_001042492); *RTEL1* (NM\_001283009; gnomAD v4.1.1 Grpmax Filtering FAF (95% confidence), including the population in which this frequency was observed (AAM = admixed American, AFAM = African/African American, FIN = Finnish; NFE= non-Finnish European; SAS = South Asian, abs = absent); CADD version 1.6 PHRED score; Splice AI delta score; AL = acceptor loss; DL = donor loss; AG = acceptor gain; DG = donor gain; ACMG-AMP classifications based on points (Tavitt et al 2020<sup>56</sup>). \*The previously reported deletion (c.1228\_1266+39del78) overlaps the one observed in our cohort but the deletion listed here is novel.

**Table 3.**
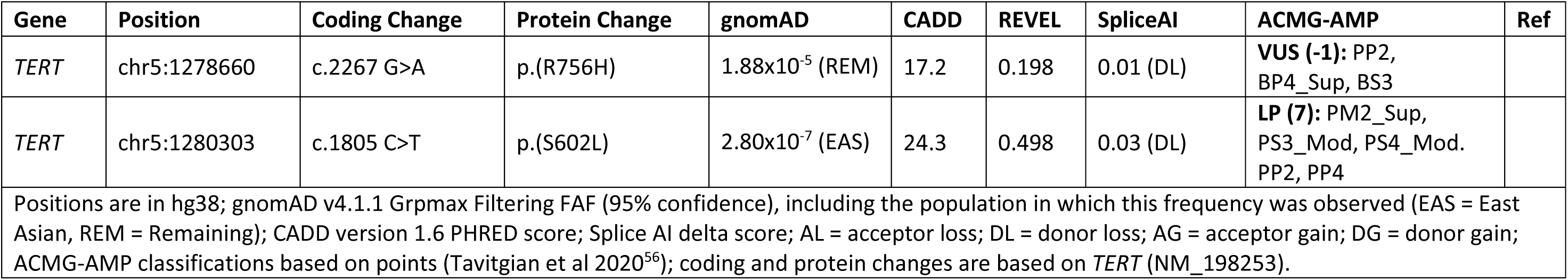
Non-segregating telomerase variants identified in the clinically confirmed PF cohort.

**Table 4.**
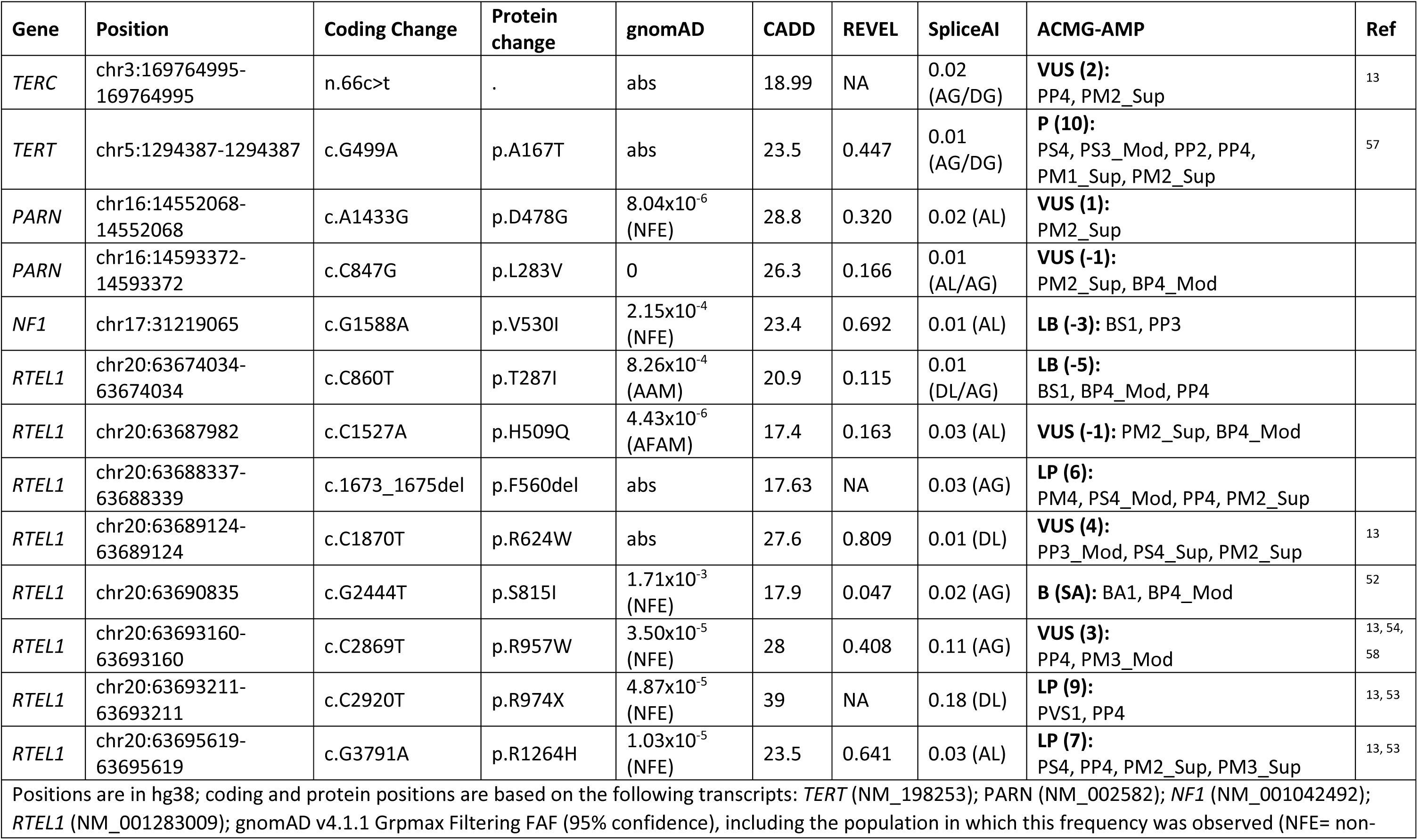
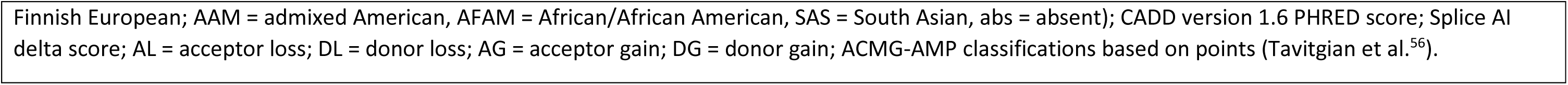
Curated variants identified in the non-familial cohort.

### Functional *TERT* assays

Four variants located in *TERT* were selected for the direct assay p.(T1111M), p.(A167T), p.(S602L) and p.(R756H). Two of these variants, p.S602L and p.R756H, were selected to explore the potential involvement in disease causation as the variant didn’t segregate with disease in the family identified. The p.(A167T) variant completely abolished both the activity and processivity of telomerase, while both the p.S602L and p.(T1111M) had defects in telomerase activity with no impact on processivity (**Figure 2a-c**). The p.(R756H) variant had no impact on telomerase activity but demonstrated approximately 85% reduction of the processivity of wild type telomerase, a result that while significant, is unlikely to be a biologically meaningful (**Figure 2c**). Controls for telomerase functional analyses are presented in supplemental data (**Figure S2**).

**Figure 2.**
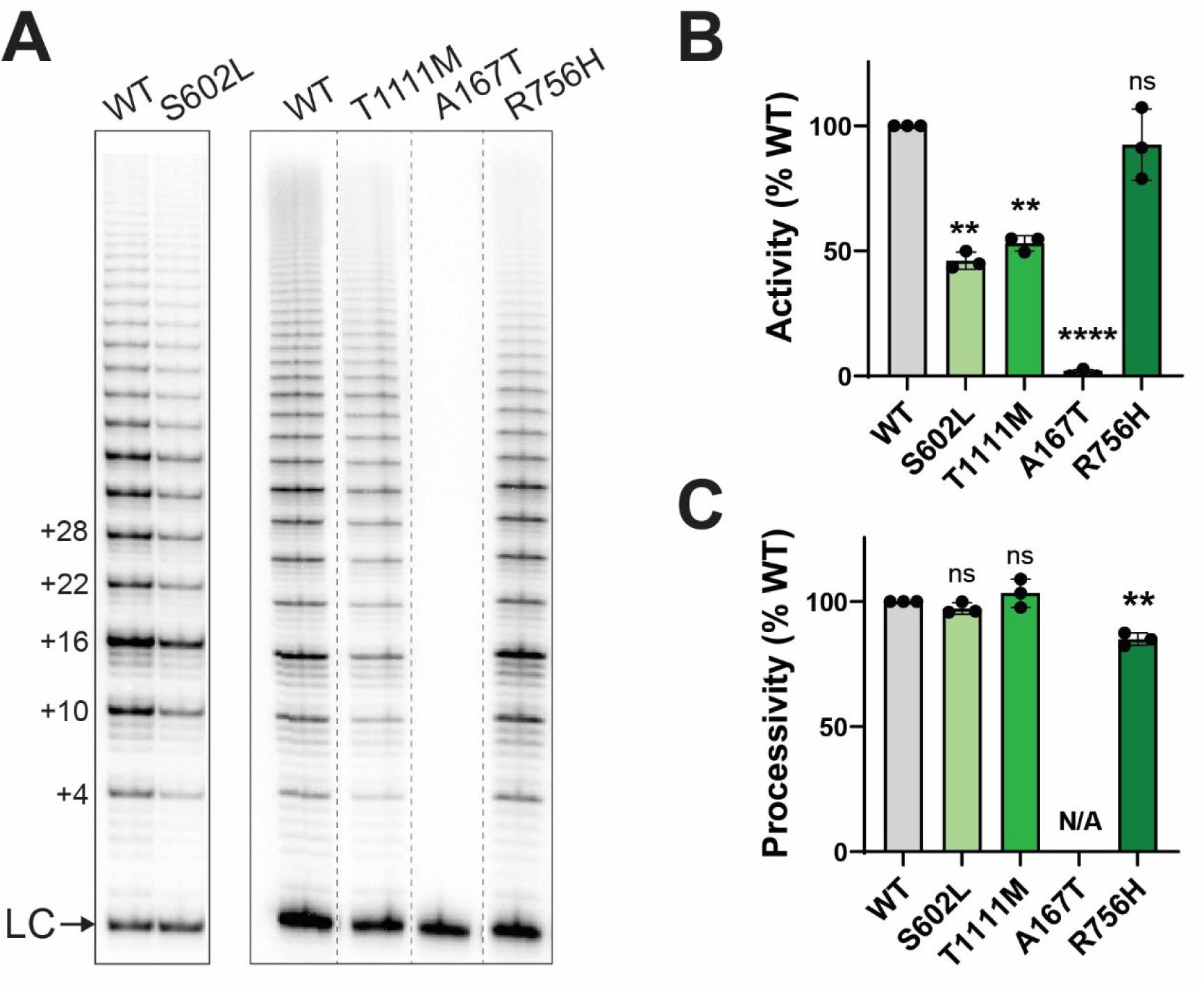
*In vitro* telomerase activity of TERT variants. **(A)** Direct primer extension assay demonstrating ability of telomerase containing the indicated TERT variants to extend a telomeric primer *in vitro*. LC = loading control; numbers at left show numbers of nucleotides added to primer. **(B)** Quantitation of telomerase activity from (A), normalised to the amount of telomerase in the reaction as determined by northern dot-blot (Supplementary Figure 1C). **(C)** Quantitation of telomerase processivity (the ability of telomerase to add multiple telomeric repeats to a single DNA molecule) from (A). Values are the mean (±SEM) of experiments from 3 independent enzyme preparations, expressed relative to WT. Significance was determined using one-sample t-test; \*\**p*<0.01, \*\*\*\**p*<0.001, ns = not significant, N/A = not applicable.

### Common genetic variant contribution to risk in rare potentially disease-causing variant carriers

Weighted PRS were generated (16 SNPs) for the familial cohort, the non-familial cohort and unscreened population controls, allowing comparison of PRS between each group (**Figure 3**). All PF cohorts showed a similar mean PRS (**Figure 3A**) and which was significantly different from the mean PRS for the control population. The individual PRS scores for the non-familial cases that were found to harbour a P/LP/VUS variant (VUS-P) were evenly distributed, with only 30.0% (n = 3/10) of these individuals having a PRS below the median of the entire sporadic cohort (0.445; **Figure 3B**). Similarly, the individual PRS scores for family members from the eleven families harbouring curated variants highlights the variable nature of the PRS in these individuals, even within families (**Figure 3C**). However, the PRS in the VUS-P variant carriers from the familial PF cohort tended to be lower, with 68.8% (n = 11/16) having a PRS below the familial PF cohort median (0.590). When the lower threshold of 0.445 was considered, representing the median PRS of the non-familial cohort, 62.5% (n = 10/16) of the familial cases still had a PRS below this value. Notably, 60.0% (n = 6/10) of the non-familial cases and 43.8% (n = 7/16) of the familial cases with VUS-P variants report as never smokers.

**Figure 3.**
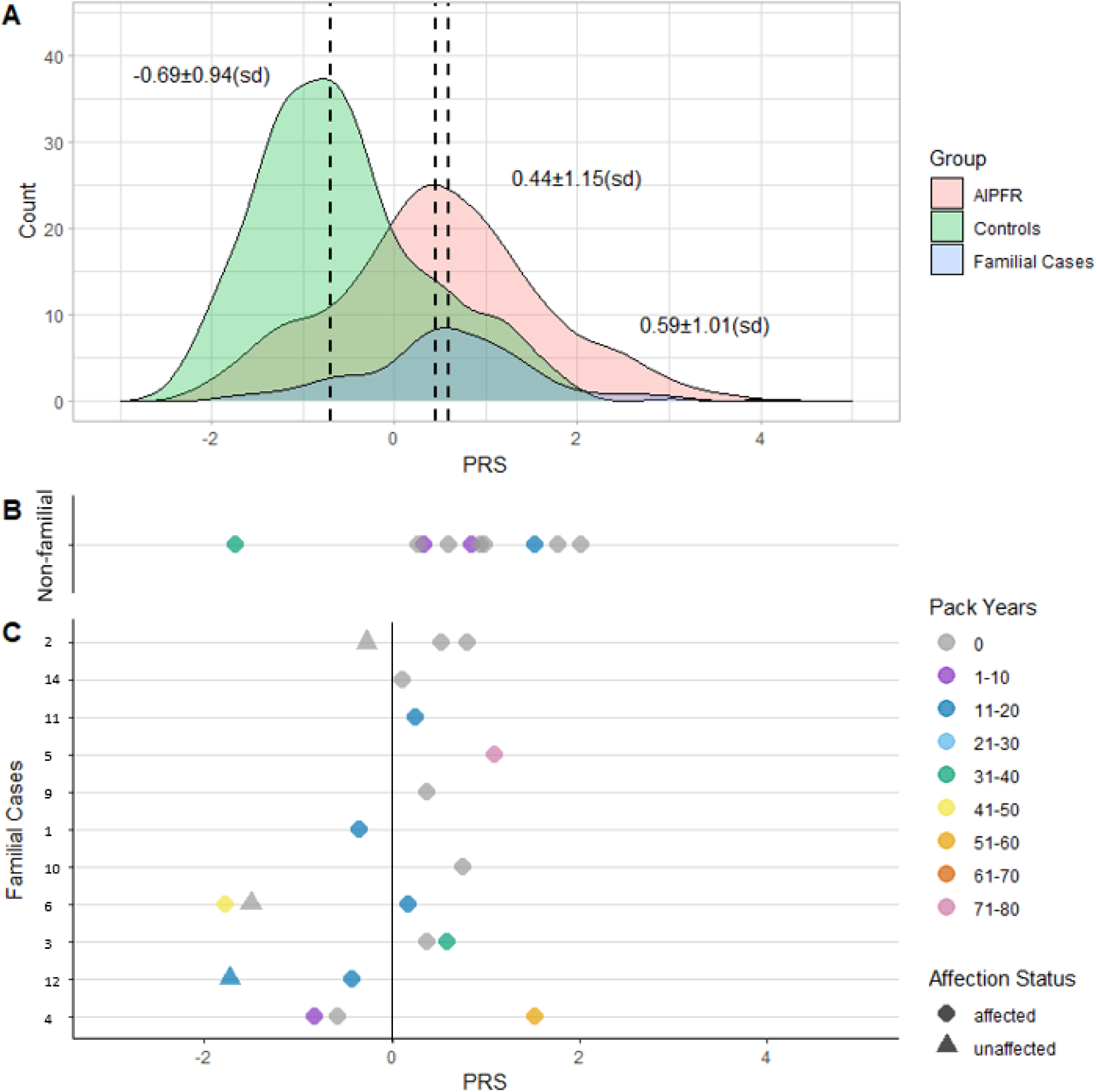
A) Histograms demonstrating the PRS distribution for population controls (green; n = 404), non-familial cases from the AIPFR (red; n = 324) and cases from the familial GRIPF cohort (blue; n=85). For each group, the median is indicated by a dotted vertical line, and the value is present along with the standard deviation (sd). B) The PRS for each individual carrying segregating variants classified as P, LP or VUS in the non-familial cohort; and C) the individuals from families (labelled vertical axis) from the familial cohort. For the families, the shape indicates affection status (with a circle indicating an affected relative and a triangle indicating an unaffected relative). The colour of the symbol indicates the self-reported smoking pack years.

Twelve of the 16 (75%) individuals with PF from the familial cohort found to have a segregating variant that was classified as VUS-P had a PRS below the median PRS for the entire familial case group (**Figure 3**). This is despite nine of the cases carrying the common risk variant at the *MUC5B* promoter. Further, in the four families with multiple cases recruited and where WGS was available, the *MUC5B* risk variant did not segregate with disease in two families (50%), and this is evident in the disparate PRS observed between the affected family members (**Figure 3**). A closer examination of family 4, where a large variation between PRS is evident, revealed that the individual with the highest burden of common risk variants was counter-intuitively diagnosed at a later age of onset despite a substantial smoking history (data not shown for privacy reasons). However, this individual is a generation older than the others in this family and all individuals with PF in this family carry a P variant in *TERT*. Therefore, the rare telomere biology variant and genetic anticipation may be a contributing factor.

### Weighted polygenic risk scores in familial and non-familial cohorts

To further examine the contribution of common risk variants to familial disease, the AIPFR cohort was divided into two groups; those self-reporting a family history (n = 41, 13.8%) and those who did not report a family history (n=284). The PRS was again significantly higher in all patient groups compared to the population controls (p < 1 x10^-5^), with no significant difference in mean PRS between AIPFR cohorts, regardless of family history, or familial cohort (**Figure 4**). The common *MUC5B* risk variant (rs35705950) was the major driver of high PRS and may be obscuring detectable differences in PRS. Removal of the MUC5B variant from the PRS resulted in no detectable statistical difference between population controls or any of patient groups (**Figure S3**), demonstrating that the PRS is largely driven by the *MUC5B* variant, consistent with previously published findings^19^.

**Figure 4.**
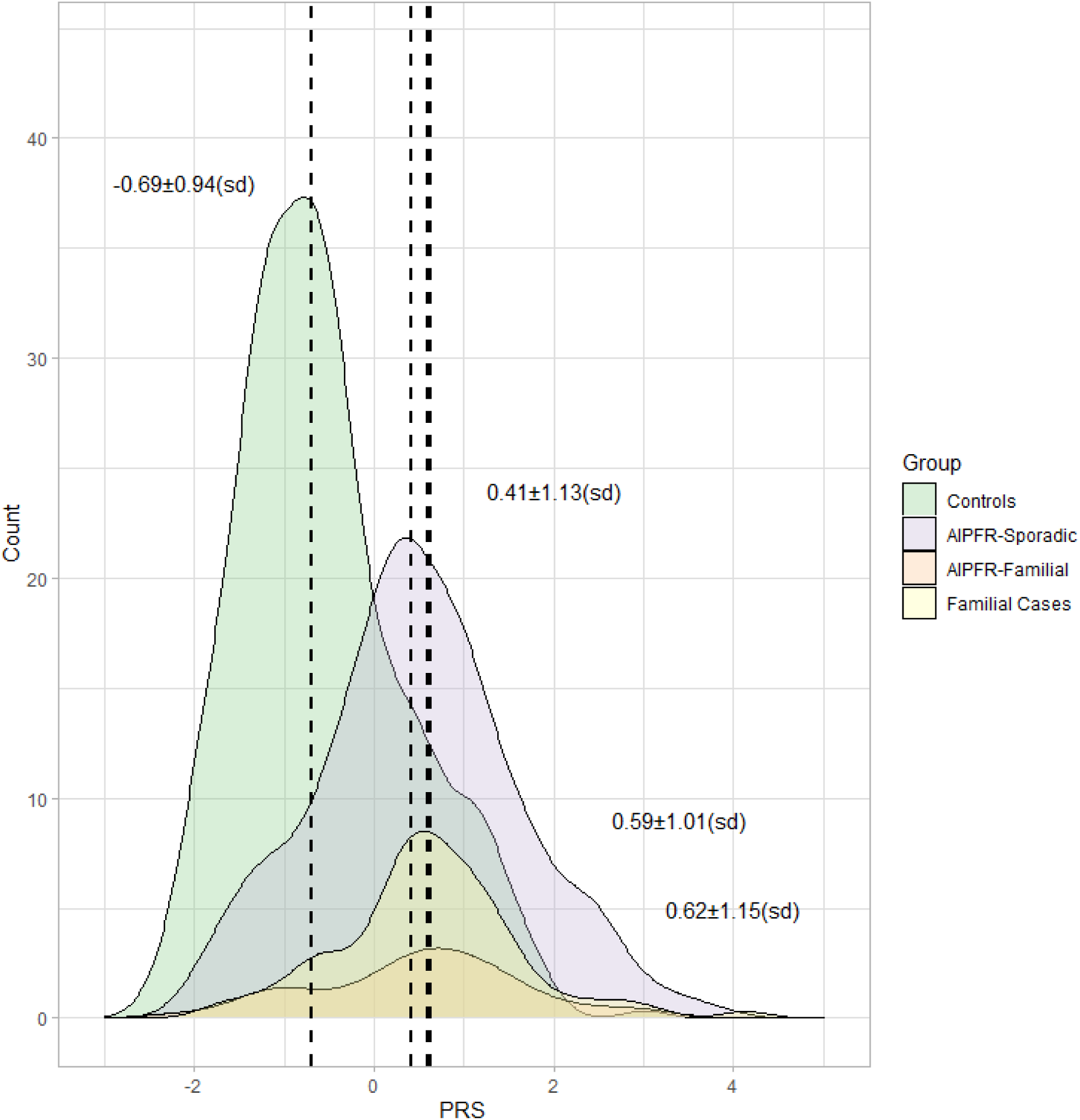
PRS were higher in all patient groups, regardless of family history (either self-reported or confirmed by a clinician), compared to population controls (1000 Genomes Project). Statistical comparison was conducted using ANOVA followed by a Tukey HSD post hoc test; p-values for all patient groups compared to the population controls was < 1 x10^-5^, while the comparisons between patient groups showed no significant difference. sd, standard deviation.

## DISCUSSION

The non-familial and familial forms of PF have been described as different ends of a spectrum of genetic risk^59^, with a higher burden of common risk variants hypothesised to explain a greater proportion of non-familial cases. Herein, we tested the hypothesis that individuals with non-familial IPF would have a higher PRS compared to the familial cases. However, there was no significant difference detected between these groups, regardless of whether family history was self-reported or clinically confirmed. We further posited that amongst familial cases where no disease-causing variants could be identified, affected status at least in a proportion of individuals, could be explained by a high burden of common risk variants, however no evidence for this was found. There was, however, evidence that families harbouring a disease-causing (P/LP) or a potentially disease-causing rare variant (VUS) were more likely to have a PRS below the median, with no clear association for the non-familial cohort. In the familial cohort, this may suggest that an underlying monogenetic cause has not yet been identified. It was also observed that PRS between cases even within the same family is highly variable. Overall, this work highlights that the genetics of IPF is highly heterogeneous, with both rare and common risk variants contributing to disease variably in different individuals regardless of family history.

This study identified fewer P/LP classified variants in known genes in the familial cohort than anticipated. Additionally, qualifying variants in *PARN* and surfactant genes were notably underrepresented in our cohort. The literature reports the frequency of *PARN* rare variants in IPF patients has been reported up to 9%^21, 60^, however none were observed in our familial cohort and only two VUSs were identified in the non-familial cohort (1.6%; n = 2/122). Additionally, across both case cohorts, only a single variant was observed in a surfactant gene (a VUS in *SFTPA2*). Consistent with published findings, the most frequently identified rare variants were identified in *TERT*^13, 15^ and *RTEL1*^13, 52^, although a large proportion of these were VUS, highlighting that even amongst established PF genes, there remains a significant gap in our understanding of variants that are likely to be causative. Fundamental challenges with curating variants, particularly in telomere biology genes, include the number of variants that are ultra rare or novel, our limited understanding of the exact function of many of the key genes, the lack of high quality of functional assays to assess gene variant impact for the majority of genes, and the limited utility of *in silico* tools predicting functionality for some genes, notably *TERT*^61^. Moreover, there are no standard rules for variant curation, and the application of the ACMG-AMP guidelines are highly variable, if used at all, resulting in a likely overrepresentation of P/LP variants in the literature. Further, as the rules are adapted overtime and more evidence becomes available, the application of criteria will change, as will some classifications. A key strength of this current study was the rigorous application of the ACMG-AMP guidelines for variant curation, including full transparency of the rules and criteria applied.

Our study highlights the importance of considering non-segregating variants in telomere biology genes as the potential cause of PF, and TBD more generally, underscoring the complex inheritance and challenges of identifying the genetic basis of disease. If the traditional family-based approach of excluding non-segregating variants based on Mendelian inheritance was applied in our cohort, a LP variant in *TERT* would have been missed and the cause of disease would not have been identified in that family. The phenomenon of a symptomatic non-carrier of the family variant has been reported previously in two separate families with TBD, where one family carried a *TERT* variant and the other a *TERC* variant^37^. If no further genetic testing or investigations were conducted such families, this would have been a missed the opportunity for targeted clinical management, although this finding is unlikely to improve risk prediction and associated genetic counselling. Beyond this however, an additional benefit of identifying the cause of the condition a family such as this is the reduction of research waste. Here, we hypothesise that the individual in this family has inherited short telomeres in the absence of the family variant and this, at a minimum, has predisposed the individual to PF. The inheritance of short telomeres in relatives without the causative family variant has also previously been reported in PF^48^ and TBD^62^. However, our data suggests that this may be a relatively common phenomenon, as even though we only considered *TERT*, non-segregating variants were identified in 8 % of multi-case families (2/25), resulting in the identification of a clinically actionable variant in one of these families (1/25; 4%). This highlights the complex inheritance of PF and TBD, even in families with apparently autosomal dominant disease.

We, among others^7–10, 63–65^, have demonstrated that up to a third of first-degree relatives of PF patients who self-reported as unaffected, were found to have pre-clinical disease suggesting that familial disease is under recognised. Our identification of clinically actionable variants in 3.7% of our non-familial cohort furthers this concept and aligns with other recent studies, prompting re-examination of the genetic similarities with familial disease^66, 67^. Further, PRS distribution was not different between apparently sporadic IPF and familial disease. Our analysis shows that the common genetic risk contribution, at least for the 16 SNPs assessed, does not differ between our IPF patients with or without a known family history. However, it is possible that this common risk burden modifies risk in either direction, even with individuals with rare variants. Common risk burden may contribute to different ages of onset or the different clinical phenotypes observed within families, although the numbers of cases harbouring LP/P variants in our study were low and limit further interpretation. There is recent evidence that a PRS for telomere length is potentially more informative in PF, although further studies are required^68^. Overall, our findings highlight that the genetic underpinnings of both familial and apparently sporadic PF are more similar than previously understood.

A limitation of this study is that the PRS was generated with only 16 validated SNPs, while the recent GWAS meta-analysis for IPF included 11,746 cases and 1,416,493 controls and identified 34 conditionally independent signals across 23 loci^17^. It is possible that generating a PRS using the larger SNP panel may reveal a more nuanced picture of common variant contribution to PRS; however, as our non-familial and familial cohorts were sourced from the same population, and the *MUC5B* variant contributes a major portion of the PRS, it is likely that our 16 SNP PRS is representative of common risk burden at least in these cohorts. This population is largely of European ancestry (self-reported); however, GWAS in other populations may return different PRS profiles, particularly those in which the *MUC5B* risk variant is rarer or absent. Of the newly identified variants not included in the current PRS, odds ratios ranged from 0.82 to 1.49, whilst the risk allele at *MUC5B* promoter variant had a frequency of 14.9% and obtained an odds ratio of 4.84 and the 15 other SNPs between 0.71 (MAF = 18.6%) and 7.82 (MAF = 0.3%)^18^. As is observed for GWAS for other complex diseases, the use of larger datasets will identify increasing number of risk variants; however, these are likely to be of diminishing effect size. Further, in terms of clinical utility and cost, a smaller number of well validated core common variants explaining the major portion of common variant contribution to disease risk is more likely to be integrated into clinical care.

The pathogenetic pathways that contribute to PF offers great promise in terms of early diagnosis and discovery of targeted treatments for PF. However, we are yet to define the relative contributions of rare and common variants, particularly in familial cases. This gap in knowledge is hampering the implementation of genetic testing for rare genetic variants in the clinical management of PF patients, as it is evident that even for those with familial disease, only a small proportion will return a definitive genetic result. This study provides a comprehensive analysis of the relative contribution of rare and common PF-associated variants in familial PF and contrasted with PF with no reported family history. We demonstrated similar PRS profiles between familial and apparently sporadic PF and identified rare disease-causing variants 9.1% of families and 3.3% of non-familial PF cases. Finally, our work highlighted the complex inheritance of PF, even in families with apparently autosomal dominant disease.

## Data Availability

All data produced in the present study are available upon reasonable request to the authors subject to the conditions for which ethics approval was obtained.

## Acknowledgements

We acknowledge the generous contribution of participants of the Genetic Research in Pulmonary Fibrosis and the Australian Idiopathic Pulmonary Fibrosis Registry without whom this research would not be possible. We also acknowledge the use of the high-performance computing facilities provided by Digital Research at the University of Tasmania. The Australian IPF Registry, an initiative of Lung Foundation Australia is supported by Foundation partners Boehringer Ingelheim, Roche Products Pty. Limited. Dr Lucas would also like to acknowledge the generous support of the Hope Fellowship, supported by Lung Foundation Australia.

## Ethics Statement

The Australian Idiopathic Pulmonary Fibrosis Registry work presented here was conducted with ethics approval obtained from the Southern Local Health District Ethics Committee with protocol numbers X11-0287 and X14-0264. Ethics approval was obtained for the Genetic Research in Pulmonary Fibrosis Study from the Southern Local Health District Ethics Committee Project ID X18-0193 & 2019/ETH06444 and the Metro North Research Ethics Committee, Queensland, Australia Project ID: 53369; HREC/2019/QPCH/53369.

