## Supplementary Material for "Mapping the common and rare variant genetic risk landscape for pulmonary fibrosis"

### 1. Supplementary Material.

List of the gene variants and gene locations and minor allele frequencies and odds ratios used for calculation of the polygenic risk scores presented.

Table S1 – Gene variants included in the PRS

| SNP | Location | Locus | Alleles | MAF | OR | Genotype method |
|---|---|---|---|---|---|---|
| rs12696304 | chr3:169763483 | *TERC (LRRC34)* | **G**/C | 27.9% | 1.31 | TaqMan |
| rs78238620 | chr3:44860894 | *KIF15(TMEM42)* | **A**/T | 5.3% | 1.58 | TaqMan |
| rs2013701 | chr4:88963935 | *FAM13A* | T/**G** | 48.7% | 0.78 | TaqMan |
| rs7725218 | chr5:1282299 | *TERT* | A/**G** | 32.5% | 0.72 | TaqMan |
| rs2076295 | chr6:7562999 | *DSP* | **G**/T | 46.9% | 1.46 | TaqMan |
| rs12699415 | chr7:1869843 | *MAD1L1* | **A**/G | 42.0% | 1.28 | TaqMan |
| rs2897075 | chr7:100032719 | *ZKSCAN1 (7q22.1)* | **T**/C | 39.1% | 1.3 | Direct seq. |
| rs28513081 | chr8:119921886 | *DEPTOR* | G/**A** | 42.8% | 0.82 | Direct seq. |
| rs537322302 | chr10:91511259 | *HECTD2* | **G**/C | 0.3% | 7.82 | TaqMan |
| rs35705950 | chr11:1219991 | *MUC5B* | G/**T** | 14.9% | 4.84 | TaqMan |
| rs9577395 | chr13:112880670 | *ATP11A* | G/**C** | 20.7% | 0.77 | TaqMan |
| rs59424629 | chr15:40428343 | *IVD* | **T**/G | 46.1% | 0.77 | Direct seq. |
| rs62023891 | chr15:85553985 | *AKAP13* | **A**/G | 30.0% | 1.27 | TaqMan |
| rs2077551 | chr17:46137522 | *MAPT(KANSL1)* | C/**T** | 18.6% | 0.71 | TaqMan |
| rs12610495 | chr19:4717660 | *DPP9* | **G**/A | 30.5% | 1.31 | TaqMan |
| rs41308092 | chr20:63693038 | *RTEL1* | **A**/G | 2.1% | 2.12 | TaqMan |

SNP = single nucleotide polymorphism; genomic location is reported according to hg38; Minor/Major alleles are listed, the effect allele is bolded; MAF = minor allele frequency; OR = odds ratio; as reported in the discovery meta-analysis from Allen et al.[1].

Table S2 – Included gene screen regions and corresponding gene (hg38).

| Chr | Start | End | Gene |
|---|---|---|---|
| chr1 | 155233452 | 155245627 | *GBA1* |
| chr2 | 85656307 | 85669741 | *SFTPB* |
| chr2 | 222565899 | 222657092 | *FARSB* |
| chr3 | 169763610 | 169766060 | *TERC* |
| chr4 | 25647011 | 25679748 | *SLC34A2* |
| chr4 | 163108133 | 163167910 | *NAF1* |
| chr5 | 1252167 | 1296068 | *TERT* |
| chr5 | 77999522 | 78295698 | *AP3B1* |
| chr5 | 139474533 | 139483758 | *STING1* |
| chr8 | 18054992 | 18085961 | *ASAH1* |
| chr8 | 22156383 | 22165479 | *SFTPC* |
| chr10 | 79554852 | 79561407 | *SFTPA2* |
| chr10 | 79609939 | 79616455 | *SFTPA1* |
| chr10 | 98415193 | 98447963 | *HPS1* |
| chr11 | 6389474 | 6395998 | *SMPD1* |
| chr11 | 59106237 | 59128412 | *FAM111B* |
| chr12 | 122470600 | 122501932 | *ZCCHC8* |
| chr14 | 22772222 | 22820796 | *SLC7A7* |
| chr14 | 24238640 | 24243674 | *TINF2* |
| chr14 | 36472288 | 36521232 | *NKX2-1* |
| chr16 | 2274881 | 2341728 | *ABCA3* |
| chr16 | 14434701 | 14631260 | *PARN* |
| chr16 | 67656512 | 67661260 | *ACD* |
| chr16 | 86509527 | 86516422 | *FOXF1* |
| chr17 | 31093927 | 31378677 | *NF1* |
| chr17 | 50055065 | 50091481 | *ITGA3* |
| chr19 | 12921479 | 12934711 | *FARSA* |
| chr20 | 63656810 | 63697253 | *RTEL1* |
| chr22 | 26442109 | 26484863 | *HPS4* |
| chr22 | 36912574 | 36941449 | *CSF2RB* |
| chrX | 1267814 | 1326218 | *CSF2RA* |
| chrX | 154761864 | 154778689 | *DKC1* |
| chrY | 1267814 | 1326218 | *CSF2RA* |

### 2. Supplementary Material

Table S3 – Gene Specific Criteria for Classification of Variants according to the ACMG/AMP Guidelines

| ACMG criterion | Strength | Specification | | | |
|---|---|---|---|---|---|
| | | *TERT* | Ref | other genes | Ref |
| **PVS1** | Sup-VStr | VStr: up to aa 1049, Str: aa1050-1132 | N | Use AutoPVS1 tool: https://autopvs1.genetics.bgi.com/ | T |
| **PS1** | Str | 1 P variant with same aa change | | | N |
| | Mod | 1 LP variant with same aa change | | | N |
| **PS2** | Sup-VStr | See et al[2] for details | | | N |
| **PS3** | Mod | Applied to direct telomerase assay (DTA) assays conducted in the laboratory of TB <75% of w/t control (with sufficient controls for moderate criteria in isolation)<br>Other published DTA assays <75% of w/t control AND telomerase extension assay (TEA) reduced OR fibroblast immortalisation impaired | N | N/A | |
| | Sup | Other published DTA assays <75% of w/t control | N | | |
| **PS4**<br>* Must meet PM2_Sup | Str | ≥ 4 published/confirmed ClinVar probands | | | N |
| | Mod | 2-3 published/confirmed ClinVar probands | | | N |
| | Sup | 1 published/confirmed ClinVar proband | | | N |
| **PM1** | Sup | Applied to the TRBD domain (aa350-601) and TEN domain (aa1-193) | N | N/A | |
| **PM2** | Sup | MAF ≤ $5.0x10^{-6}$, based on the frequency of the known LP and P variants Arg671Trp (LP, 8 alleles in gnomAD, MAF = $4.5x10^{-6}$) and Val694Met (P, 6 alleles in gnomAD, MAF = $1.9x10^{-6}$) | | Dominant/semi-dominant inheritance: MAF ≤ $1.25x10^{-5}$<br>Recessive inheritance: MAF ≤ $1.0x10^{-3}$ | |
| **PM3** | Mod | N/A | | *RTEL1* only if recessive inheritance observed<br>*In trans* with LP/P variant | T |
| | Sup | | | *RTEL1* only if recessive inheritance observed<br>Homozygous | T |
| **PM4** | Mod | In-frame indel in TRBD and TEN domains | N | In-frame indel (not applied with PVS1) | T |
| **PM5** | Sup-Str | See et al[2] for details | | | N |

| | | | | | |
|---|---|---|---|---|---|
| **PM6** | Sup-VStr | See et al[2] for details | | | N |
| **PP1** | Str | ≥7 meioses | | | N |
| | Mod | 5-6 meioses | | | N |
| | Sup | 3-4 meioses | | | N |
| **PP2** | Sup | Apply to all TERT variants unless BA1 BS1 is met | N | N/A | T |
| **PP3**<br>* missense | Str | N/A | | REVEL score ≥ 0.932 | P<br>W |
| | Mod | N/A | | REVEL score 0.773-0.931 | |
| | Sup | REVEL > 0.55 | N | REVEL score 0.644-0.772 OR<br>SpliceAI ≥ 0.2 | |
| **PP4** | Sup | Apply if 2 or more phenotypic features of TBD | | | N |
| **PP5** | | N/A | | | |
| **BA1** | SA | MAF ≥ 0.005 | N | Dominant/semi-dominant inheritance: MAF ≥ 0.00125<br>Recessive inheritance: MAF ≥ 0.05 | |
| **BS1** | Str | MAF ≥ 0.000625 | N | Dominant/semi-dominant inheritance: MAF ≥ 0.000125<br>Recessive inheritance: MAF ≥ 0.01 | |
| **BS2** | | N/A | | | |
| **BS3** | Sup | DTA activity > 75% of wild-type controls | N | N/A | |
| **BS4** | | N/A | | | |
| **BP1** | | N/A | | | |
| **BP2** | | N/A | | | |
| **BP3** | | N/A | | | |
| **BP4** | Str | N/A | | REVEL score ≤ 0.016 | P |
| | Mod | N/A | | REVEL score 0.017-0.183 | |
| | Sup | REVEL < 0.43 | N | REVEL score 0.184-0.290 | |
| **BP5** | | N/A | | | |
| **BP6** | | N/A | | | |
| **BP7** | Sup | PhyloP <0.1 (and SpliceAI <0.75) | N | N/A | T |

- MAF = Grpmax Filtering AF (95% confidence) in gnomAD v4.1.0
- N = Nelson et al 2023 (PMID: 36496180)
- T = Thompson et al 2025 (PMID: 39279436)
- P = Pejaver et al 2022 (PMID: 36413997)

- W = Walker et al 2023 (PMID: 37352859)

3. Supplementary Material

Estimates of Relatedness in Individuals from the GRIPF Cohort

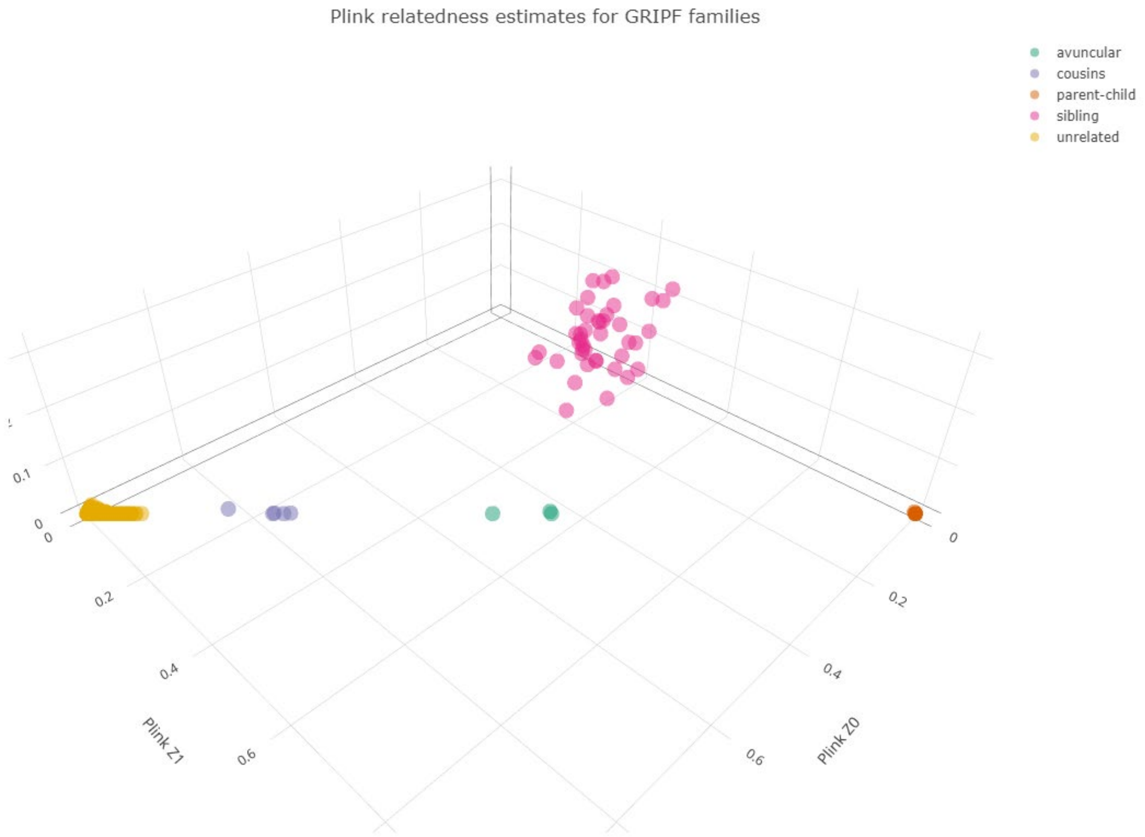

Figure S1 – Estimates of Relatedness in Individuals from the clinically confirmed GRIPF Cohort. Relationship testing confirmed all reported relationships and confirmed no cryptic relationships between families. An interactive version of this plot is available here: https://chart-studio.plotly.com/~semlucas/17.

4. Supplementary Data: Variants Excluded from Curation in the AIPFR Cohort

In the AIPFR Cohort of those individuals with no reported family history 23 individuals harboured a heterozygous variant in genes previously only known to be associated with autosomal disease. Of these three individuals were found to harbour two qualifying variants located in a gene known to cause autosomal recessive disease. This included one individual with two variants in SLC34A2 and two individuals with biallelic variants in AP3B1. One individual carried two separate variants in the SLC34A2 gene (NM_006424): p.P73T and p.L414F. Biallelic variants in this gene are associated with pulmonary alveolar microlithiasis which is rarely associated with PF, but has been reported to mimic IPF[3]. However, both variants were classified as LB and therefore these variants are not considered to be the potential cause of disease in this individual. Additionally, two individuals harboured two qualifying variants in the AP3B1 (NM_003664) gene. Pathogenic homozygous or compound heterozygous variants in this gene cause Hermansky-Pudlak syndrome 2, a severe syndrome characterised by platelet defects, oculocutaneous albinism, immunodeficiency and commonly pulmonary fibrosis[4]. Both individuals carried the p.V999M variant, while they additionally carried p.I357V and p.A400P, respectively. Notably, the p.V999M variant was observed in an additional individual with self-reported familial PF that carried two variants in this gene, as well as two families in the clinically confirmed familial PF cohort, though was not prioritised in this group due to the lack of a second variant in the gene as well as incomplete segregation. The self-reported health information and the clinician questionnaire was reviewed for all three individuals that carried the p.V999M variant and a second AP3B1 variant, however none of the other clinical features of Hermansky–Pudlak syndrome were reported. All three AP3B1 variants were classified as LB and therefore there was insufficient evidence to assess causation in these individuals. All five variants were subsequently excluded from further analyses (Table S4).

Table S4 – Qualifying variants identified in the non-familial cohort that were excluded as potentially disease causing due to the lack of pathogenicity

| **Gene** | **Position*** | **Coding Change** | **Protein change** | **Cohort** | **gnomAD** | **CADD** | **REVEL** | **ACMG-AMP** |
|---|---|---|---|---|---|---|---|---|
| *SLC34A2* | chr4:25662809 | c.C217A | p.P73T | NF | $6.88x10^{-6}$ (NFE) | 19.9 | 0.18 | **LB (-2):** BP4_Mod |
| *SLC34A2* | chr4:25674321 | c.G1242C | p.L414F | NF | $4.96x10^{-3}$ (NFE) | 22.8 | 0.14 | **LB (-2):** BP4_Mod |
| *AP3B1* | chr5:78015426 | c.C3115A | p.Q1039K | SRF | $2.92x10^{-6}$ (NFE) | 22.5 | 0.161 | **VUS (-1):** PM2_Sup, BP4_Mod |
| *AP3B1* | chr5:78015546 | c.G2995A | p.V999M | SRF and NF | $8.28x10^{-3}$ (NFE) | 15.3 | 0.072 | **LB (-2):** BP4_Mod |
| *AP3B1* | chr5:78165642 | c.G1198C | p.A400P | NF | $2.33x10^{-3}$ (NFE) | 24.9 | 0.173 | **LB (-2):** BP4_Mod |

| | | | | | | | | |
|---|---|---|---|---|---|---|---|---|
| *AP3B1* | chr5:78175810 | c.A1069G | p.I357V | NF | $2.04x10^{-3}$ (NFE) | 20.9 | 0.122 | **LB (-2):** BP4_Mod |
| *Positions are in hg38; Cohorts included self-reported familial (SRF), familial (F) and non-familial (NF); gnomAD v4.1.1 Grpmax Filtering FAF (95% confidence), including the population in which this frequency was observed (NFE= non-Finnish European); CADD version 1.6 PHRED score. SLC34A2: NM_006424, AP3B1: NM_003664 | | | | | | | | |

5. Supplementary Data

Controls for telomerase functional analyses

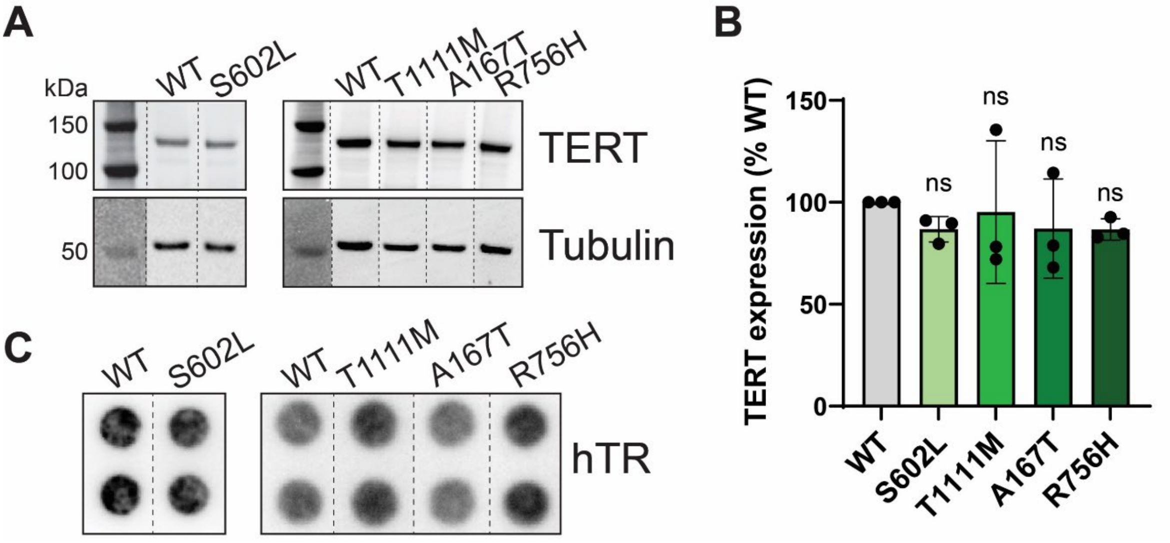

Figure S2: Controls for telomerase functional analyses. (A) Western blots showing TERT expression from the indicated plasmids transfected into HEK293T cells, and tubulin as a loading control. (B) Quantitation of TERT expression from (A); values are the mean (±SEM) of three blots from three independent enzyme preparations, normalised to tubulin, expressed relative to WT, with significance determined using one-sample t-test; ns = not significant. (C) Northern dot-blot showing amounts of hTR after immunoprecipitation of TERT from HEK293T lysates after transfection of the indicated plasmids; each sample loaded in duplicate.

6. Supplementary Data

PRS Scores (excluding the MUC5B Variant) generated for AIPFR and GRIPF datasets compared to the 1000 Genomes dataset (1KG)

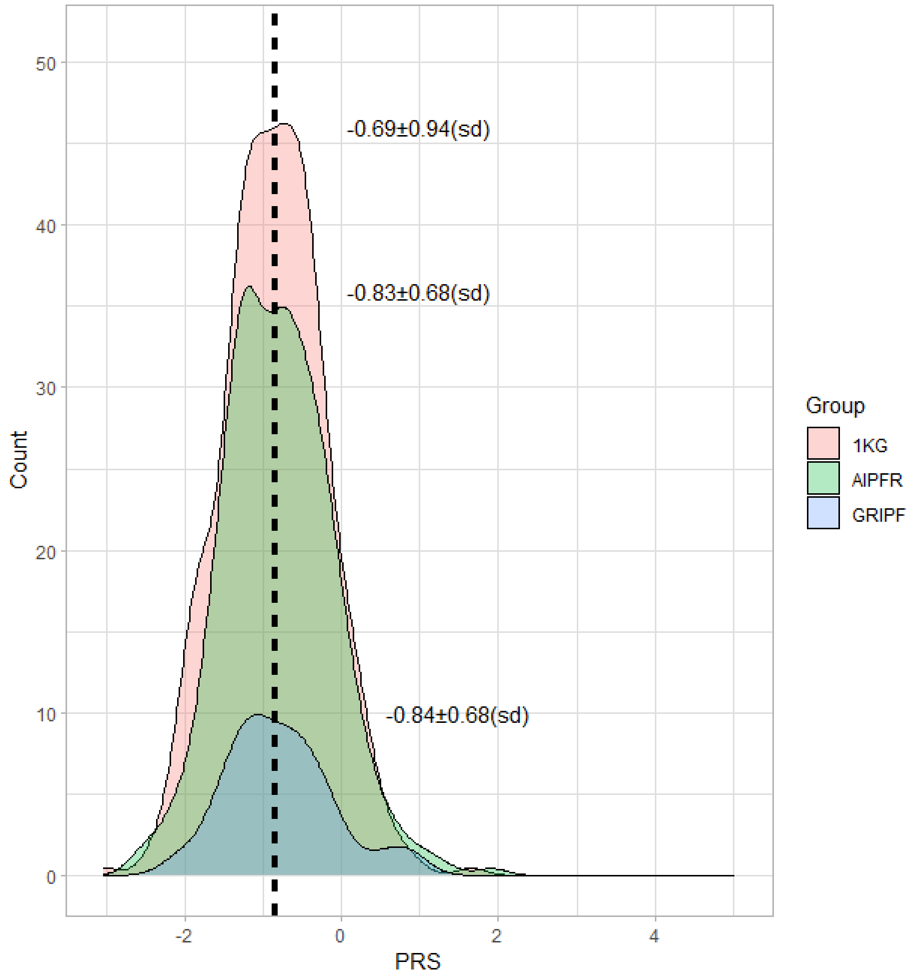

Figure S3 - PRS scores when excluding the MUC5B promoter variant. Histograms demonstrating the PRS distribution for population controls from the 1000 genomes project (1KG; red; n = 404), non-familial cases from the AIPFR (green; n = 324) and cases from the clinically confirmed familial GRIPF cohort (blue; n=85). For each group, the median is indicated by a dotted vertical line and the value is present along with the standard deviation (sd). No significant difference between groups was observed.
